# First-in-human case report: AAV9-hGAA gene therapy for a patient with infantile-onset Pompe disease

**DOI:** 10.1101/2022.12.22.22283398

**Authors:** Xiuwei Ma, Jun Li, Xiaodong Wang, Wenhao Ma, Jianhua Wang, Ruijie Gu, Zhiming Zhu, Yongxia Wang, Ying Du, Juan Xu, Fang He, Xiao Yang, Sheng Zhang, Lina Zhu, Qiuping Li, Hui Xiong, Xiaobing Wu, Zhichun Feng

## Abstract

**Background:** The classic infantile-onset Pompe disease (IOPD) is characterized by cardiac hypertrophy, respiratory insufficiency, and rapidly progressive muscle weakness due to the acid alpha-glucosidase (GAA) deficiency. Enzyme replacement therapy (ERT) is the current approach for IOPD, but it entails several limitations. Aiming to overcome the limited efficiency of ERT, we developed adeno-associated virus (AAV) gene therapy for IOPD patients.

**Method:** One IOPD patient received a single intravenous dose of GC301, a recombinant adeno-associated virus 9 (rAAV9) expressing the human GAA (rAAV-hGAA). During the follow-up, safety was accessed by the physical examinations, cardiac and laboratory evaluations. GAA activity, the titers of serum antibodies to AAV9 and GAA, and motor development were monitored regularly.

**Result:** The infant showed significant improvements in motor milestones. The GAA enzyme activity increased to the normal range. The cardiac function improved notably.

**Conclusion:** In patient with IOPD, a single intravenous AAV9-hGAA gene therapy improved the clinical outcomes remarkably. The trial is still ongoing, the safety of this gene therapy and the long-term clinical benefit remain to be monitored for months and years to come.

## Introduction

Pompe disease, also known as glycogen-storage disease type II or acid maltase deficiency, is a metabolic disorder^1^. It is an autosomal recessive trait characterized by acid alpha-glucosidase (GAA) deficiency leading to lysosomal glycogen storage. The classic infantile-onset Pompe disease (IOPD) is characterized by cardiac hypertrophy, respiratory insufficiency, and rapidly progressive muscle weakness^1^. The infants will die before the age of 1 year without typical treatment^2^. Enzyme replacement therapy (ERT) is the standard of care to treat Pompe disease now. But this treatment entails several limitations, for example, the short drug half-life and an antibody response resulting in reduced efficacy. AAV-associated gene therapy provides a way to prolong the efficacy of GAA enzymatic activity. In this investigator-initiated clinical trial, an AAV-associated gene-drug was used to treat an IOPD infant by intravenous infusion.

## Method

### The patient

A baby within two months of the birth, the outpatient examination found cyanosis of lips and motor delay. Echocardiography showed hypertrophic cardiomyopathy with increases in inter-ventricular septum thickness in diastole (IVSD) and left ventricular mass (LVM). Genetic tests revealed two compound heterozygous variants, one of which was a maternal frameshift variant (c.258dup, p.Asn87Glnfs*9), and the other was a paternal stop-gain variant (c.2284G > T, p.Glu762*) (Figure S1A), while the GAA activity was 0.14 μmol/L/h (reference value is 1.46 - 20.34 μmol/L/h) (Figure 2A). The infant started ERT immediately after diagnosis. The patient received Myozyme (alfa-glucosidase, 20 mg/kg) treatment twice before administration of GC301 (Figure 1).

**Figure 1.**
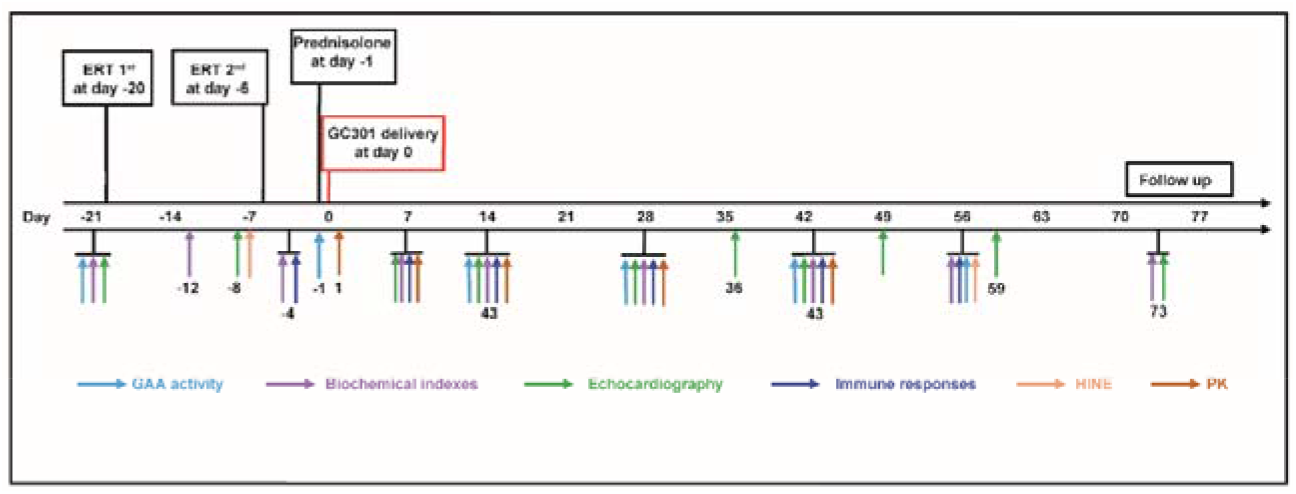
The graphic overview of the clinical trial including the initial time of prednisolone, GC301 administration (Day 0), timepoints of sampling for PK, GAA enzymatic activity check, laboratory and physical tests, etc.

**Figure 2.**
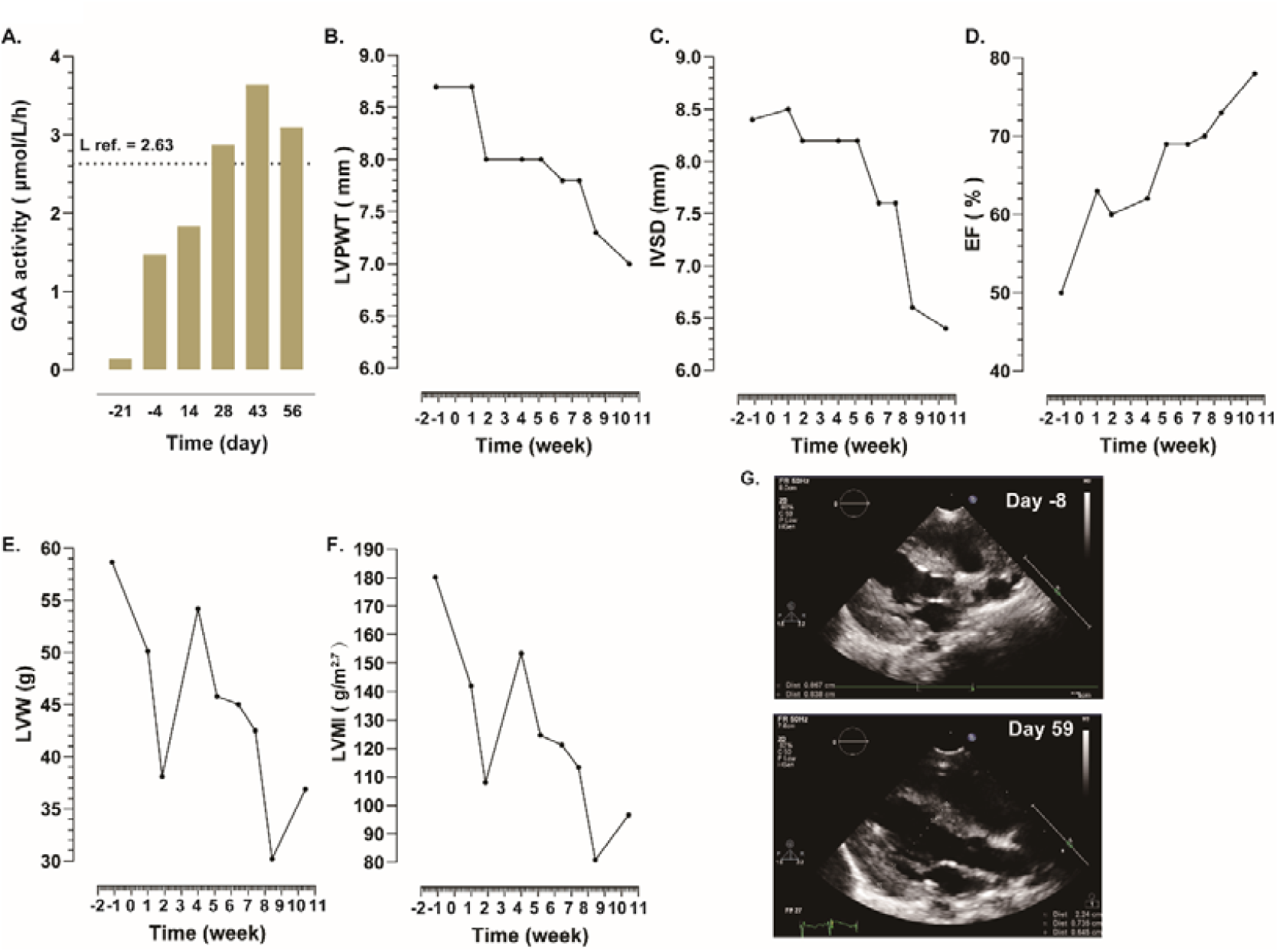
The changes in GAA activity and cardiac function during the course of clinical treatment. The changes of GAA activity(A), LVPWT(B), IVSD(C), EF(D), LVW(E), and LVMI(F) during the whole process of treatment by now. Ultrasound image map of before (day −8) and after (day 59) GC301 treatment was shown in G.

### GC301

GC301 is a one-single dose gene drug based on rAAV9 in which the codon-optimized human GAA was transcribed under the control of the constitutive promoter. GMP-grade GC301 was manufactured and supplied by Genecradle Therapeutics Inc..

### Study procedure

In this study, the day with GC301 infusion was noted as day 0. The IOPD patient received a single intravenous dose of 1.2 × 10^14^ vector genomes per kilogram bodyweight (vg/kg) of GC301. The day before GC301 delivery, the infant received prophylactic oral prednisolone at 2 mg/kg/day last for two days, then a reduced dose of 1 mg/kg/day on the third day. This dose (1 mg/kg/day) lasted 4 weeks and tapered from the fifth week until the stop. The total prednisolone course was 58 days. Drug safety and efficacy have been observed for 10 weeks post-treatment for the moment. The whole treatment and monitoring process were shown in Figure 1.

### Clinical evaluation

According to the study protocol, the safety of GC301 was accessed by the reported adverse events (AE) based on the vital signs, physical examinations, cardiac and laboratory evaluation. GAA activity was monitored every 14 days. The titers of serum antibodies to AAV9 and GAA were detected and analyzed simultaneously. Motor development was assessed by the Hammersmith Infant Neurological Examination (HINE).

## Result

### Clinical outcomes

The infant showed significant motor improvement with better head-control and reached some other developmental milestones for 4-6 months infant including raising his head when lying face down and rocking on his stomach. The patient was assessed by the HINE before GC301 delivery (the −4^th^ day) and on the 56^th^ Day. The HINE scores were 54 and 69 respectively (Table 1).

**Table 1.**
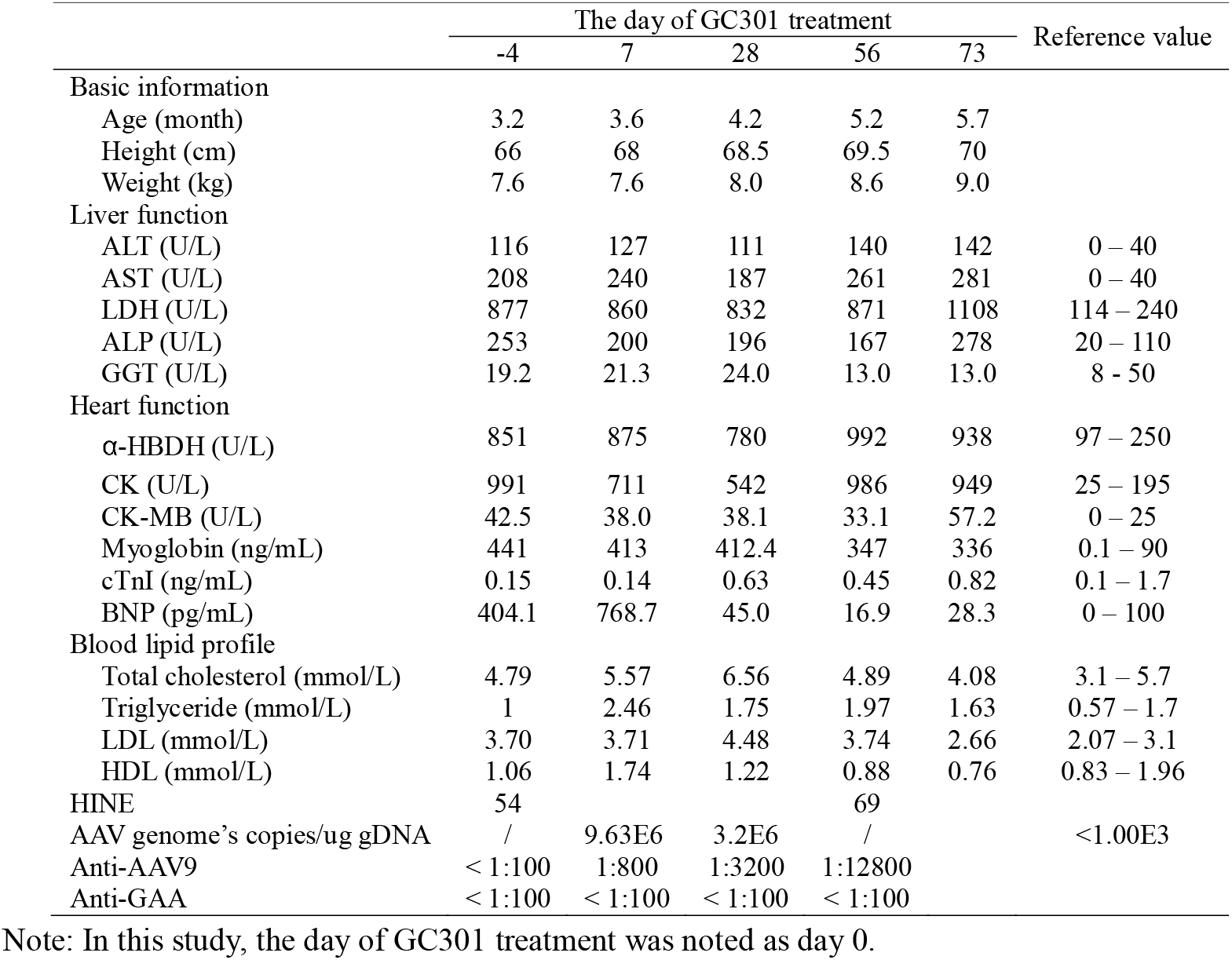
The biochemical indexes during the trial

### GAA enzyme activity

As the main index, GAA activity plays an important role in assessing the clinical benefit from the therapy. The levels of plasma GAA enzyme activity were analyzed during the whole therapeutic process including the ERT and GC301 injection. The GAA activity was lower than the low reference (the value was 1.47 μmol/L/h, reference is 2.63 - 21.69 μmol/L/h) before GC301 delivery (the −4^th^ day). The GAA activity began to increase, and the value was 2.90 μmol/L/h at the end of the 4^th^ week (Figure 2A). Since then, the value had remained within the normal range. The value of GAA activity was 3.64 μmol/L/h on the 43^rd^ day and 3.1 μmol/L/h on the 56^th^ day.

### Cardiac response

We measured the left ventricular posterior wall thickness (LVPWT), interventricular end septal diastole (IVSD), and left ventricular ejection fraction (EF) by echocardiography. Echo reveals that the left ventricular wall thickness and mass in this infant increased at the time of diagnosis. Cardiac hypertrophy was alleviated after the GC301 delivery. The LVPWT of the infant reduced from 8.7 mm to 7.0 mm on the 7^th^ day and the 73^rd^ day respectively (Figure 2B). The IVSD decreased from 8.5 mm on the 7^th^ day to 6.4 mm on the 73^rd^ day (Figure 2C). The value of EF was 78% on the 73^rd^ day while the value was only 63% on the 7^th^ day (Figure 2D). The left ventricular weight (LVW) declined to 30.20 g on the 59^th^ day from 50.13 g on the 7^th^ day (Figure 2E). The left ventricular mass index (LVMI) declined from 142.00 g/m^2.7^ on the 7^th^ day to 80.80 g/m^2.7^ on the 59^th^ day (Figure 2F). The more intuitive changes in ventricular wall thickness can be seen in the ultrasound image of day −8 and day 59 (Figure 2G).

### Changes in biochemical indexes

The infant serum protein changed followed by GC301 treatment. The serum alanine transaminase (ALT), aspartate transaminase (AST), lactate dehydrogenase (LDH), alkaline phosphatase (ALP), hydroxybutyrate dehydrogenase (α-HBDH), creatine kinase (CK), creatine kinase-MB (CK-MB), and myoglobin always stayed higher than the high reference value (Table 1). The cardiac troponin I (cTnI) was always kept in the normal ranges. The brain natriuretic peptide (BNP) level decreased into the range of reference values on the 28^th^ day (Table 1). The total cholesterol level increased to the highest on the 28^th^ day (the value is 6.56 mmol/L), then fall into the normal ranges (the value was 4.89 mmol/L on the 56^th^ day and the value was 4.08 mmol/L on the 73^rd^ day) (Table 1). The level of triglyceride showed the same trend as total cholesterol, that is the triglyceride level raised to the highest on the 7^th^ day (the value is 2.46 mmol/L) and then fall into the normal ranges on the 73^rd^ day (the value is 1.63 mmol/L) (Table 1). The level of low-density lipoprotein (LDL) and high-density lipoprotein (HDL) showed similar change trends (Table 1). The level of LDL raised and then fall into the normal ranges. The value of HDL always kept into the normal ranges, but the latest result declined below the low reference value.

## Discussion

Pompe disease is a multi-systemic condition with manifestations in the cardiac, skeletal muscles, and nervous system caused by GAA enzymatic deficiency^3^. ERT was approved by the FDA in 2006 and is the only available treatment for Pompe disease by now. Albeit the ERT improved IOPD clinical outcomes, multiple factors including short enzyme turnover time in the blood requiring lifelong usage, low uptake efficiency in skeletal muscle cells, and preferential uptake in the liver leading to enzyme clearance, provide the limited efficiency of ERT^4,5^. By contrast, a single dose of an AAV-based drug may solve the aforementioned problems and achieve comparable and/or even better clinical effects. In this study, we evaluated the safety and efficacy of GC301, a recombinant adeno-associated virus expressing GAA in a Chinese pediatric patient with IOPD. Especially as we are aware, this is the first gene therapy trial on IOPD patient in the world. To date, no SAE was observed and favorable results suggested GC301 therapeutic efficacy.

GAA enzyme level is a pertinent indicator in assessing the efficacy of gene therapy. In this trial, the GAA activity elevated from the 2^nd^ week after GC301 infusion and raised higher than the lower threshold of clinical reference value on the 4^th^ week and thereafter. We postulated that GC301 could express the transgene GAA rapidly and improve the GAA enzyme level in the blood within two weeks. GAA proteins secreted into the peripheral blood from the targeted tissue cells and those from peripheral blood leucocytes expressing exogenous GAA could contribute to the elevated enzyme level in the blood^6,7^. However, the anti-GAA antibody level was kept under the detective limits throughout the course, suggesting that the transduced target cells were not eradicated by cellular immunity.

The patient received GC301 infusion 5 days after the second dose of ERT. Peak plasma recombinant human GAA alglucosidase alfa (Myozyme) levels were present directly after infusion, and GAA enzyme levels in the blood declined and cleared within several hours subsequently^8,9^. GAA enzyme activity climbed higher than the lower threshold of the normal range (2.63-21.69 μmol/L/h) and reached 3.64 μmol/L/h in our patient on the 6^th^ week after GC301 infusion. This enzyme activity was even higher than that of his normal mother (2.96 μmol/L/h) who is the heterozygous carrier of GAA gene variant c.258dup (p. Asn87Glnfs*9). Different from the impulsive impact of ERT in the targeted tissue, elevated and sustained GAA enzyme levels in the blood should be attributed to the successful GC301 transduction of target cells and continuous production of therapeutic transgene product providing sustained levels in transduced cells and/or secreted levels in plasma for cross-correction.

Among the overall clinical changes, cardiac function improved most significantly. Within 8 weeks after GC301 intravenous infusion, there was an increased EF and a reduction in left ventricular mass suggesting cardiac correction because of GC301. Compared with other gene therapy trials and ERT for IOPD, faster cardiac improvement was observed in this study^10-12^. Previous study demonstrated that ten IOPD patients achieved normal LVM (mean Z score = 0.9±0.8) at week 104 after ERT^12^, whereas our patient’s LVM Z score was 1.05 at 60^th^ day post GC301 infusion. In this study, we used the AAV9, which has favorable expression in the heart, at an infusion dose of 1.2×10^14^ vg/kg to treat this infant. GC301 concentration in the blood dropped about 4-logs compared to two hours post-treatment on the 7^th^ day (1.39E10 VS. 9.63E6). This result is consistent with our PK data in non-human primates (unpublished data). We speculated that the rapid clearance of vector DNA in plasma was associated with the fast transduction of the gene into target cells due to the AAV9 heart and liver tropism^13^.

During the follow-up, no severe adverse event (SAE) was observed. We found an asymptomatic elevation on total cholesterol and triglyceride, which only occurred transiently and recovered without any intervention. This observation is different from other AAV gene therapy trials where the clinically significant elevations of liver enzymes were mostly observed in SAE and AE post-treatment^14-16^. Increases in serum aminotransferase (ALT and AST) concentrations were noted at the 1^st^, 4^th^, 8^th^ and 10^th^ weeks post-IV injection of the AAV vector. These elevations were usually associated with the high titers of anti-AAV antibodies^16^. The ALT and AST levels at baseline for this patient raised to 3∼5 times of upper limit of normal due to the natural course of the disease. However, ALT and AST levels kept relatively stable despite the high titers of the anti-AAV9 antibody (1:12800 in the 8^th^ week) after GC301 infusion. Other liver function laboratory evaluations, for example, Gamma-glutamyl transferase (GGT) were normal during the trial, indicating that liver function was not affected post-treatment. Although we used a relatively high vector dose, GC301 showed no harm to liver.

In summary, current results on motor development and cardiac response demonstrated that GC301 gene therapy improved the IOPD patient clinical outcomes remarkably. In addition, the patient’s vital signs and laboratory evaluation results showed that the treatment was safe. Although our data so far suggested the safety and efficacy of GC301, the observation period was just 10 weeks after GC301 injection, the trial is still ongoing, and the long-term clinical benefit remains to be monitored for months and years to come.

## Data Availability

All data produced in the present study are available upon reasonable request to the authors.

## Author Contributions

Zhichun Feng, Xiaobing Wu and Hui Xiong conceived and designed the clinical trial. Xiuwei Ma, Xiaodong Wang, Ruijie Gu, Jianhua Wang, Hui Xiong, Yongxia Wang, and Wenhao Ma executed the research. Jun Li, Wenhao Ma, Yongxia Wang, Fang He, and Ruijie Gu collected and interpret the data. Xiuwei Ma, Jun Li, Xiaodong Wang, Ying Du, Wenhao Ma, and Zhiming Zhu contributed to the data analysis. Xiuwei Ma, Jun Li, and Xiao Dong Wang drafted the original manuscript, and Xiaobing Wu, Qiuping Li, and Zhichun Feng edited the manuscript. Juan Xu contributed to the vector production quality control. Sheng Zhang, Lina Zhu, and Xiao Yang contributed to patient follow-up and data collection.

## Declaration of interests

We declare no competing interests.

## Ethics

This trial was approved by the ethics committee of the seventh medical center of PLA General Hospital (S2022-004-01), and human genetic resource administration of China [2022] SLCJ2122. Clinical trial registered at Chictr.org.cn (ChiCTR2200063229) and Clinicaltrials.gov (NCT05567627). The legal guardian of the infant voluntarily consented to take part in the research study and authorized the use and disclosure of patient’s genetic and clinical information in connection with the study.

## Fund

This project was supported by the Natural Science Foundation of China Youth Fund (32100640).

## Acknowledgments

We thank the patient and his parents for participating in this study.

## Figure legend

**Figure S1.**
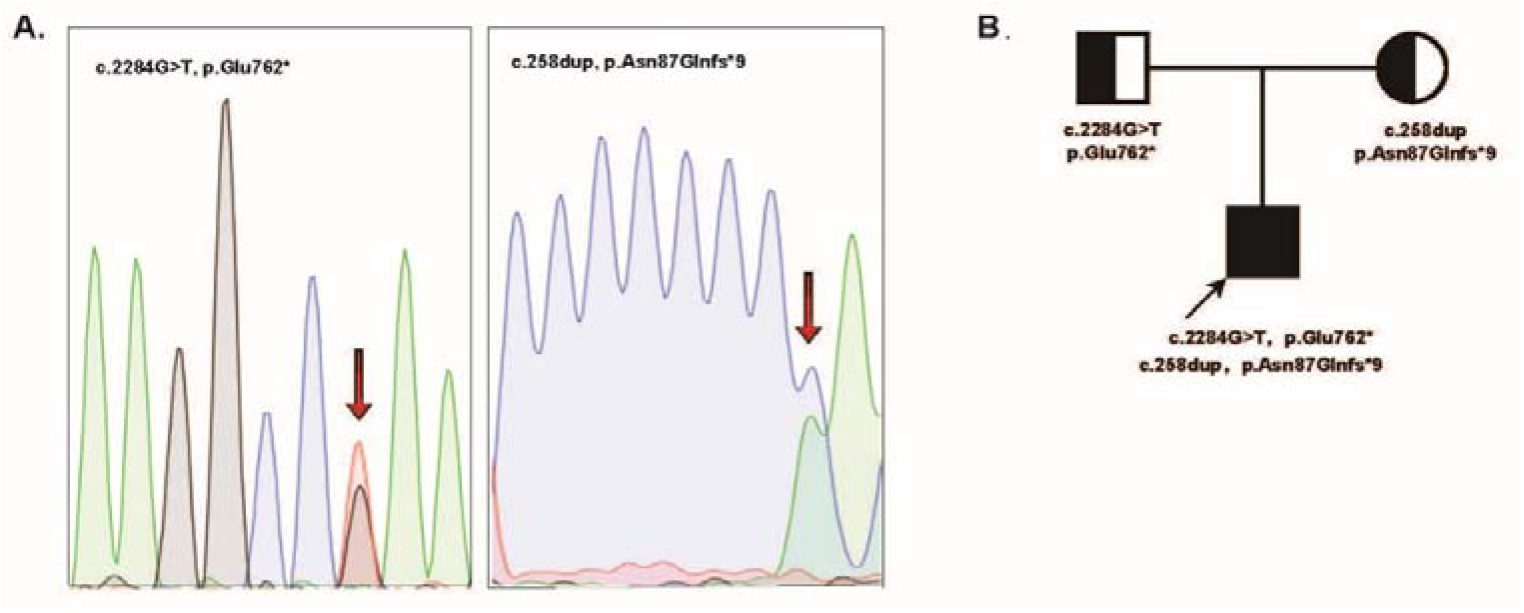
Sanger sequencing revealed compound heterozygous truncating variants c.258dup (p. Asn87Glnfs*9), c.2284G>T (p. Glu762*) in the patient’s *GAA* gene (A). The pedigree map illustrated the variants in the patient (arrow) and his parents (B).

## Notes

### Competing Interest Statement

The authors have declared no competing interest.

### Clinical Trial

NCT05567627

### Author Declarations

This trial was approved by the ethics committee of the seventh medical center of PLA General Hospital (S2022-004-01)

